# Systematic Screening of Prior Medication Use Reveals Protective Associations Against Schizophrenia Linked to *Toxoplasma gondii*-Targeting Agents

**DOI:** 10.1101/2025.02.14.25322266

**Authors:** Ariel Israel, Abraham Weizman, Sarah Israel, Joshua Stokar, Shai Ashkenazi, Shlomo Vinker, Eli Magen, Eugene Merzon

**Author notes:** **Corresponding author:** Ariel Israel, Leumit Research Institute, Sprinzak 23, Tel-Aviv 6473817, Israel.

## Abstract

Schizophrenia (SCZ) is a severe psychiatric disorder with a complex and poorly understood etiology. Previous studies have linked *Toxoplasma gondii* infection to SCZ, yet its clinical relevance remains unclear. In this study, we systematically screened all medication purchases and medical diagnoses recorded from 10 years to 30 days before SCZ diagnosis to identify factors associated with altered SCZ risk. Using electronic health records spanning over two decades from a national health provider, we retrospectively compared 3,273 SCZ patients with 32,730 controls matched for demographics, socioeconomic status, ethnicity, and year of enrollment. Statistical tests were adjusted for multiple comparisons using a false discovery rate (FDR), with significant associations (FDR<10%) further evaluated in multivariable models to account for residual confounding. Although the analysis included all medication classes, agents targeting *T. gondii* emerged as the most strongly protective, most notably atovaquone/proguanil, clindamycin, and ophthalmic fluoroquinolones. Protective effects were also observed for agents known to reduce neuroinflammation, such as nonsteroidal anti-inflammatory drugs (NSAIDs), particularly COX-2 inhibitors. In contrast, increased SCZ risk was associated with several other compounds, including neomycin, tramadol, and desmopressin. These findings were validated in the U.S.-based TriNetX research network (>120M individuals), with prospective survival analyses of propensity score-matched cohorts, selected by historical medication exposure, confirming key associations with high statistical significance (P<0.0001). These findings support the hypothesis that *T. gondii* induced neuroinflammation may contribute to SCZ pathogenesis and suggest that targeted antiparasitic and anti-inflammatory interventions could offer therapeutic potential. Further studies are warranted to explore the clinical relevance of these associations.

## Introduction

Schizophrenia (SCZ) is a chronic psychiatric disorder affecting ∼1% of the global population, ^1^ marked by hallucinations, delusions, and cognitive deficits.^2^ Although genetic studies highlight substantial heritability, environmental factors are increasingly recognized as pivotal in its pathogenesis. Prior research has linked *Toxoplasma gondii* infection to SCZ,*^3^* yet causality remains unconfirmed. Here, we analyze over 20 years of electronic health record (EHR) data to explore whether prior medical treatments influence SCZ risk. Using EHRs from Leumit Health Services (LHS), a national healthcare provider, we compared 3,273 SCZ patients with 32,730 matched controls to identify distinct patterns in past treatments, diagnoses, and laboratory results. Our goal was to uncover significant associations shedding light on risk and protective factors for SCZ. If *T. gondii* contributes causally, treatments effective against the parasite should correlate with reduced SCZ incidence.

## Methods

### Study design

The first stage of this research was conducted as a retrospective cohort study in Leumit Health Services (LHS), one of the four national health providers in Israel, providing comprehensive healthcare services to approximately 730,000 members. All Israeli citizens are entitled for comprehensive health insurance and receive a standardized package of health services and medications, as defined by the national “Health Basket” committee. LHS operates a centralized EHR system, with over two decades of meticulously maintained information on patient demographics, medical diagnoses, healthcare encounters, laboratory test results, and records of prescribed and purchased medications. Diagnoses are documented during medical encounters by the treating physicians using the International Classification of Diseases, Ninth Revision (ICD-9). Diagnoses can be marked as chronic when they pertain to a chronic condition, and these can be updated or closed by the treating physicians during subsequent patient encounters. The reliability of these chronic diagnosis records in our registry has been previously validated, demonstrating high accuracy.^4^

Eligibility for inclusion in the study was defined as any past or current LHS member with at least three years of LHS membership between years 2003 and 2024. Data extraction was carried out from the LHS central data warehouse, encompassing diagnoses, results of laboratory tests, and medication purchases recorded up to December 31, 2024. Data extraction was automated using structured query language (SQL) and Python scripts to retrieve relevant patient information from the LHS data warehouse. Socioeconomic status was determined using geocoded residential addresses and categorized on a scale from 1 (lowest) to 20 (highest) based on the Points Location Intelligence® database, which highly correlates with socioeconomic measures provided by the Israeli Central Bureau of Statistics. Geodemographic classification of patients into the general population, Ultra-Orthodox Jewish population, and Arab population was performed using validated methodologies established in prior studies. Demographic data, laboratory test results, and physical measures, including body mass index (BMI) and blood pressure (BP), were extracted from the EHR. Smoking status and physical measures were documented by treating physicians or nurses during routine medical visits. The most recent available record prior to the index date was used for analysis if available, otherwise the first record after the index date was taken. While annual check-ups are not mandatory for all LHS members, national quality measures ensure regular documentation of smoking status, BMI, BP by healthcare providers.

### Cohort definition

The study cohort consists of eligible SCZ patients diagnosed for the first time between the ages of 13 and 40, alongside a control group matched at a 10:1 ratio, selected from LHS members who had no SCZ diagnosis. To avoid misclassification due to overlapping conditions, individuals with a diagnosis indicative of a cerebrovascular accident were excluded from both groups of the study.

SCZ patients were identified based on the presence of an ICD-10 “F20” coded diagnosis of Schizophrenia recorded in the EHRs. A patient was classified as having SCZ if the diagnosis was recorded by a psychiatrist or, when recorded by a non-psychiatrist physician in the list of chronic conditions, it had not been subsequently closed and was accompanied by documented purchases of antipsychotic medication for at least five months.

The index date was defined as the earliest recorded SCZ diagnosis in the EHR or the date of the first antipsychotic medication purchase, if the medication was obtained up to one year before the SCZ diagnosis. To ensure that the date of SCZ onset was accurately determined and that medical history prior to disease onset could be extracted from the EHR, we required the first diagnosis of SCZ to occur at least two full years after LHS membership enrollment. Control individuals were matched to SCZ patients based on gender, ethnic group (categorized as ultra-Orthodox, of Arab descent, or the general population), socioeconomic status (SES) category, and year of initial enrollment in LHS. For each SCZ patient, ten control individuals were selected who met these criteria. Among potential control candidates, those with the closest birth date to the SCZ patient were chosen, ensuring that no individual was duplicated within the cohort.

### Data processing

Data were extracted from electronic health records and processed for analysis using scripts developed by the Leumit Research Institute in Python 3.11, utilizing the Pandas library and T-SQL queries. Before analysis, patient data were deidentified, with patient ID numbers replaced by study-specific internal identifiers.

Medication consumption was analyzed based on the purchase records classified according to the medication Anatomical Therapeutic Chemical (ATC) classification and route of administration and covered the period from 10 years before the index date to 30 days prior.

Medical conditions were assessed based on physician visits that occurred up to 30 days before the index date. The analysis used a curated list of approximately 200 common medical conditions, compiled by the Leumit Research Institute and identified by their corresponding ICD-9 diagnosis codes or laboratory tests.

### Statistical analyses

Statistical analyses were conducted using R version 4.4. Unless otherwise specified, Fisher’s exact test was used to compare categorical variables, and the two-tailed, two-sample t-test was applied to compare continuous variables between groups. For these analyses, we report odds ratios (ORs) with 95% confidence intervals (CIs). Conditional logistic regression models were employed to evaluate associations between the SCZ group and independent variables, including medication use and diagnostic records across matched groups. These models adjusted for potential overlaps between medications and medical conditions, as well as confounders included as covariates: age (in years), gender, smoking status, socioeconomic status (SES) category, number of prior physician visits, history of pregnancy, and healthcare worker status. For each variable in the regression models, we report the adjusted odds ratio (aOR) with its 95% CI. Sensitivity analyses in the LHS cohort demonstrated that the leading associations, including Ato/Pro, remained statistically significant even when earlier cut-off points - several years prior to the index date - were used to define exposure windows.

### TriNetX cohort

Validation analyses were conducted using TriNetX, a global federated health research network that provides access to EHRs, including diagnoses, procedures, medications, laboratory values, and genomic information, from large healthcare organizations (HCOs). The validation studies were performed within the US Collaborative Network, which comprises 70 HCOs, and over 120 million patients. While retrospective in terms of data collection, the TriNetX design was prospective in structure, with cohorts defined based on exposure and followed forward in time for incident SCZ.

We utilized the “compare outcomes” analysis framework, focusing on individuals currently aged between 13 and 40 to ensure cohort homogeneity. Case individuals were defined as those who received the medication of interest between ages 10 and 30, while control individuals were those who had an ambulatory visit at the same age, which served as the index date in both groups.

To ensure balance at medication initiation date, propensity score matching (PSM) was applied based on age, ethnicity, race, BMI presence, and smoking status. SCZ occurrence was compared between the two cohorts using a Cox proportional hazards model, with follow-up starting after the first occurrence of the index event. We did not limit the follow-up end time.

### Ethics Statement

The Institutional Review Board (IRB) of Leumit Health Services gave ethical approval for this work (LEU-0020-24), with a waiver of informed consent, since data were analyzed retrospectively and anonymously.

## Results

Table 1 compares demographic and clinical characteristics of SCZ and control patients. Matched variables, as expected, showed similarity between groups, with both having comparable age and gender distributions (76.1% male, mean age 24.2 ± 7.2 years). SCZ patients exhibited higher smoking rates (OR=1.43, 95% CI [1.31–1.55]), slightly elevated diastolic blood pressure (72.1 ± 9.3 vs. 71.2 ± 9.2 mmHg, SMD=0.098), and reduced total T3 levels. They also had increased prevalence of subclinical hypothyroidism (TSH>4.5 mIU/L, OR=2.00 [1.69–2.36]), vitamin D deficiency (OR=1.38 [1.18–1.61]), and folic acid deficiency (OR=1.41 [1.20–1.64]). These results are consistent with prior studies on metabolic and endocrine changes in SCZ.^5–8^

**Table 1.** Demographic and clinical characteristics of the study cohort at index date.

|  |  | **SCZ**<br>**N=3,273** | **Control**<br>**N=32,730** | **P-**<br>**value** | **Odds Ratio** | **SMD** |
| --- | --- | --- | --- | --- | --- | --- |
| Gender | Female | 782 (23.9 %) | 7,820 (23.9 %) | 1.000 | 1 |  |
|  | Male | 2,491 (76.1 %) | 24,910 (76.1 %) | 1.000 | 1 |  |
| Age (years) |  | 24.2 ± 7.2 | 24.2 ± 7.2 | 0.979 |  | 0.001 |
| Age category | 10-18 | 914 (27.93 %) | 8,978 (27.43 %) | 0.983 | 1.03 [0.95–1.11] |  |
|  | 19-29 | 1,537 (46.96 %) | 15,495 (47.34 %) | 0.985 | 0.98 [0.92–1.06] |  |
|  | 30-39 | 772 (25.1%) | 7,749 (25.2 %) | 0.931 | 1.00 [0.91–1.08] |  |
|  | 40 | 50 (1.53 %) | 508 (1.55 %) | 1.000 | 0.98 [0.72–1.32] |  |
| Weight (kg) |  | 68.1 ± 19.1 | 68.4 ± 19.3 | 0.511 |  | -0.012 |
|  | missing | 73 (2.23 %) | 2,280 (6.97 %) |  |  |  |
| Height (cm) |  | 168 ± 12 | 168 ± 13 | 0.041 |  | -0.038 |
|  | missing | 122 (3.73 %) | 3,248 (9.92 %) |  |  |  |
| Body Mass Index (kg/m2) |  | 24.0 ± 5.6 | 24.0 ± 5.4 | 0.857 |  | 0.003 |
|  | missing | 110 (3.36 %) | 3,007 (9.19 %) |  |  |  |
| BP systolic (mmHg) |  | 117 ± 17 | 117 ± 14 | 0.004 |  | 0.055 |
|  | missing | 171 (5.22 %) | 2,595 (7.93 %) |  |  |  |
| BP diastolic (mmHg) |  | 72.1 ± 9.3 | 71.2 ± 9.2 | <0.001 |  | 0.098 |
|  | missing | 171 (5.22 %) | 2,611 (7.98 %) |  |  |  |
| Smoking status | Non-smoker | 2,040 (67.93 %) | 20,893 (74.67 %) | <0.001 | 0.72 [0.66–0.78] |  |
|  | Past smoker | 23 (0.77 %) | 316 (1.13 %) | 0.079 | 0.68 [0.42–1.03] |  |
|  | Smoker | 940 (31.30 %) | 6,771 (24.20 %) | <0.001 | 1.43 [1.31–1.55] |  |
|  | missing | 270 (8.25 %) | 4,750 (14.51 %) | <0.001 | 0.53 [0.46–0.60] |  |
| eGFR MDRD (mL/min/1.73m2) |  | 129 ± 48 | 130 ± 56 | 0.422 |  | -0.017 |
|  | missing | 809 (24.7 %) | 9,689 (29.6 %) |  |  |  |
| Glucose (mg/dL) |  | 88.5 ± 16.0 | 88.4 ± 14.7 | 0.705 |  | 0.080 |
|  | missing | 919 (28.1 %) | 10,572 (32.3 %) |  |  |  |
| Total T3 (triiodothyronine) (nmol/L) |  | 1.78 ± 0.42 | 2.00 ± 0.54 | <0.001 |  | -0.409 |
|  | < 1.2 nmol/L | 15 (4.04 %) | 34 (1.44 %) | 0.002 | 2.88 [1.44–5.51] |  |
|  | missing | 2,902 (88.7 %) | 30,367 (92.8 %) |  |  |  |
| Free T3 (pmol/L) |  | 4.92 ± 1.33 | 4.99 ± 1.27 | 0.036 |  | -0.049 |
|  | < 3.5 pmol/L | 13 (3.77 %) | 111 (2.12 %) | 0.006 | 1.81 [1.17–2.24] |  |
|  | missing | 2,451 (74.9 %) | 27,496 (84.0 %) |  |  |  |
| TSH (Thyroid Stimulating Hormone) (mIU/L) |  | 2.30 ± 2.26 | 2.05 ± 2.02 | <0.001 |  | 0.123 |
|  | > 4.5 mIU/L | 187 (6.15 %) | 872 (3.17 %) | <0.001 | 2.00 [1.69–2.36] |  |
|  | missing | 234 (7.15 %) | 5,195 (15.87 %) |  |  |  |
| Vitamin D (ng/mL) |  | 18.9 ± 8.9 | 20.1 ± 9.1 | <0.001 |  | -0.137 |
|  | < 30 ng/mL | 1,935 (90.29 %) | 14,939 (87.11 %) | <0.001 | 1.38 [1.18–1.61] |  |
|  | missing | 1,130 (34.5 %) | 15,581 (47.6 %) |  |  |  |
| Folic Acid (ng/mL) |  | 6.61 ± 3.85 | 6.83 ± 3.70 | <0.001 |  | -0.059 |
|  | < 2.8 ng/mL | 217 (9.22 %) | 1,254 (6.74 %) | <0.001 | 1.41 [1.20–1.64] |  |
|  | missing | 920 (28.1 %) | 14,120 (43.1 %) |  |  |  |

SMD: Standardized Mean Difference (Cohen's D)

### Medications associated with SCZ risk

We investigated which classes of prior medication use were most associated with the risk of SCZ. For each medication classes present in our health system, we assessed their usage in both the SCZ and control groups over the 10 years preceding the index date. A significantly higher usage rate in the SCZ group compared to controls suggests that the medication may be associated with an increased risk of SCZ. Conversely, a significantly lower usage rate in the SCZ group may indicate a potential protective effect against SCZ occurrence.

Table 2 presents the medications with the most significant differences in past usage rates between the SCZ and control groups over the decade preceding the index date, categorized by therapeutic class. Statistical significance was assessed using Fisher’s exact test, and only medications with a false discovery rate (FDR) below 10% were retained.

**Table 2.**
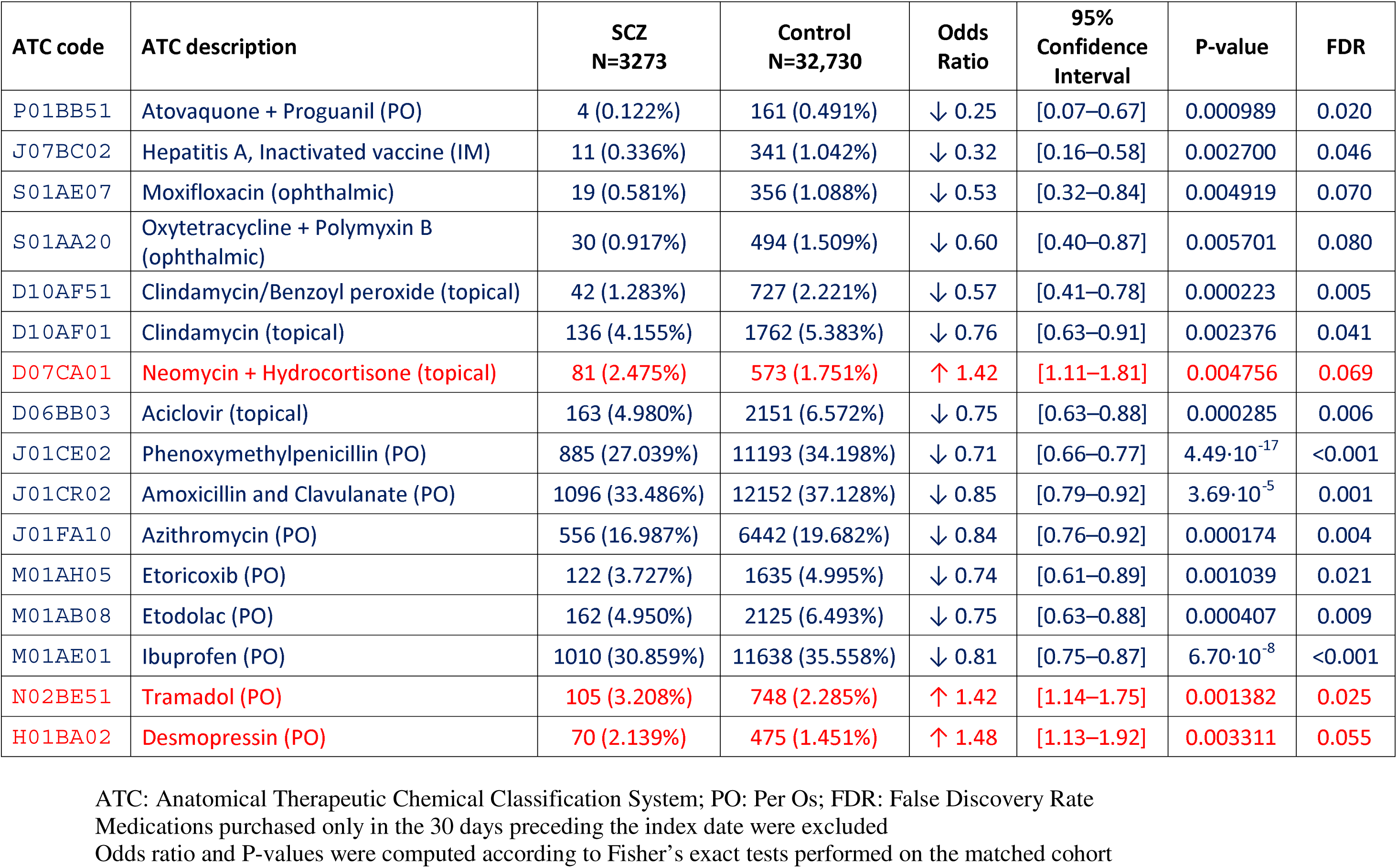
Comparison of medication purchases in the 10 years to 30 days before the index date in the SCZ and control groups.

| ATC code | ATC description | SCZ<br>N=3273 | Control<br>N=32,730 | Odds<br>Ratio | 95%<br>Confidence<br>Interval | P-value | FDR |
| --- | --- | --- | --- | --- | --- | --- | --- |
| P01BB51 | Atovaquone + Proguanil (PO) | 4 (0.122%) | 161 (0.491%) | ↓ 0.25 | [0.07–0.67] | 0.000989 | 0.020 |
| J07BC02 | Hepatitis A, Inactivated vaccine (IM) | 11 (0.336%) | 341 (1.042%) | ↓ 0.32 | [0.16–0.58] | 0.002700 | 0.046 |
| S01AE07 | Moxifloxacin (ophthalmic) | 19 (0.581%) | 356 (1.088%) | ↓ 0.53 | [0.32–0.84] | 0.004919 | 0.070 |
| S01AA20 | Oxytetracycline + Polymyxin B (ophthalmic) | 30 (0.917%) | 494 (1.509%) | ↓ 0.60 | [0.40–0.87] | 0.005701 | 0.080 |
| D10AF51 | Clindamycin/Benzoyl peroxide (topical) | 42 (1.283%) | 727 (2.221%) | ↓ 0.57 | [0.41–0.78] | 0.000223 | 0.005 |
| D10AF01 | Clindamycin (topical) | 136 (4.155%) | 1762 (5.383%) | ↓ 0.76 | [0.63–0.91] | 0.002376 | 0.041 |
| D07CA01 | Neomycin + Hydrocortisone (topical) | 81 (2.475%) | 573 (1.751%) | ↑ 1.42 | [1.11–1.81] | 0.004756 | 0.069 |
| D06BB03 | Aciclovir (topical) | 163 (4.980%) | 2151 (6.572%) | ↓ 0.75 | [0.63–0.88] | 0.000285 | 0.006 |
| J01CE02 | Phenoxymethylpenicillin (PO) | 885 (27.039%) | 11193 (34.198%) | ↓ 0.71 | [0.66–0.77] | 4.49·10<sup>-17</sup> | <0.001 |
| J01CR02 | Amoxicillin and Clavulanate (PO) | 1096 (33.486%) | 12152 (37.128%) | ↓ 0.85 | [0.79–0.92] | 3.69·10<sup>-5</sup> | 0.001 |
| J01FA10 | Azithromycin (PO) | 556 (16.987%) | 6442 (19.682%) | ↓ 0.84 | [0.76–0.92] | 0.000174 | 0.004 |
| M01AH05 | Etoricoxib (PO) | 122 (3.727%) | 1635 (4.995%) | ↓ 0.74 | [0.61–0.89] | 0.001039 | 0.021 |
| M01AB08 | Etodolac (PO) | 162 (4.950%) | 2125 (6.493%) | ↓ 0.75 | [0.63–0.88] | 0.000407 | 0.009 |
| M01AE01 | Ibuprofen (PO) | 1010 (30.859%) | 11638 (35.558%) | ↓ 0.81 | [0.75–0.87] | 6.70·10<sup>-8</sup> | <0.001 |
| N02BE51 | Tramadol (PO) | 105 (3.208%) | 748 (2.285%) | ↑ 1.42 | [1.14–1.75] | 0.001382 | 0.025 |
| H01BA02 | Desmopressin (PO) | 70 (2.139%) | 475 (1.451%) | ↑ 1.48 | [1.13–1.92] | 0.003311 | 0.055 |

ATC: Anatomical Therapeutic Chemical Classification System; PO: Per Os; FDR: False Discovery Rate
Medications purchased only in the 30 days preceding the index date were excluded
Odds ratio and P-values were computed according to Fisher's exact tests performed on the matched cohort

We further conducted a multivariable regression analysis to adjust for potential confounders, including the overlapping effects of qualifying medications, as well as age, gender, socioeconomic status, smoking history, prior pregnancy, and healthcare utilization. For selected cases, we also included medications from the same class or with similar indications to evaluate whether the observed associations reflected broader therapeutic effects or were influenced by confounding due to overlapping treatments.

Figure 1 presents a forest plot of the adjusted multivariable logistic regression results. Atovaquone/proguanil (Ato/Pro) (aOR=0.26, 95% CI: [0.09–0.70]) and hepatitis A vaccine (aOR=0.31 [0.17–0.57]) showed strong protective effects. Ophthalmic antimicrobials, including moxifloxacin (aOR=0.53 [0.33–0.85]) and oxytetracycline/polymyxin B (aOR=0.61 [0.42–0.89]), were also protective. Topical clindamycin, alone or with benzoyl peroxide, reduced SCZ risk, whereas benzoyl peroxide alone increased it, indicating clindamycin-specific protection. Conversely, neomycin-based topical treatments heightened SCZ risk, particularly the ointment formulation (aOR=1.54 [1.21–1.96]), an effect not explained by associated antifungal or corticosteroid components. Other topical antibiotics (mupirocin, gentamicin, chloramphenicol) similarly increased risk. Among oral antibiotics, penicillin, amoxicillin/clavulanate, and azithromycin showed moderate protection. NSAIDs, especially COX-2 inhibitors (etoricoxib, etodolac, ibuprofen), reduced SCZ risk, while tramadol (aOR=1.62 [1.30–2.00]) and desmopressin (aOR=2.04 [1.49–2.80]) were linked to increased risk.

**Figure 1:**
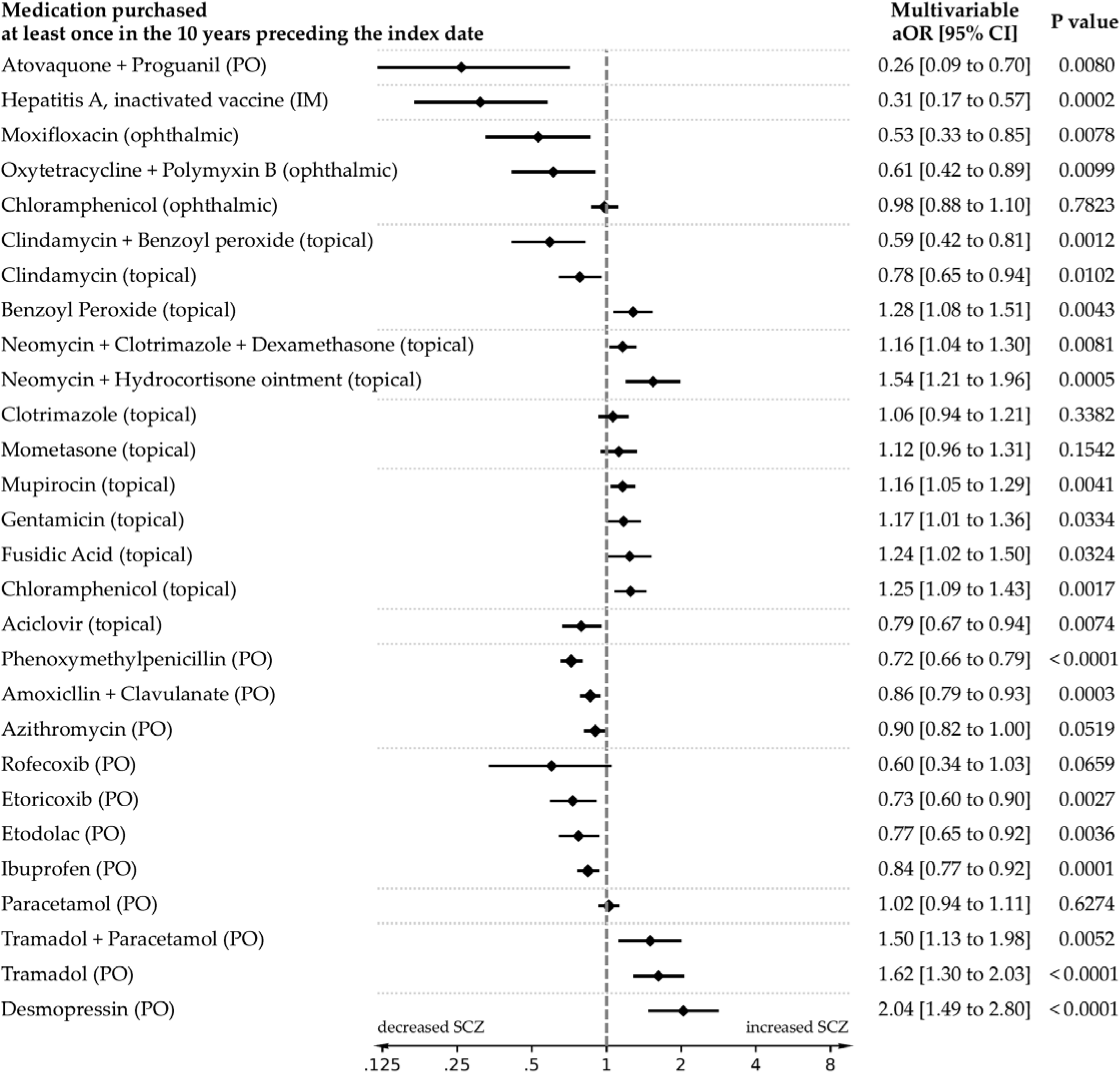
Forest plot of schizophrenia risk according to medication purchase history in the 10 years preceding the index date. The multivariable conditional logistic regression model included all the listed medications and is adjusted for age (years), gender, smoking status, SES category, number of past physician visits, previous pregnancy, and health worker status aOR: Adjusted Odds Ratio; PO: Per Os; IM: Intramuscular

### Medical conditions associated with SCZ risk

We assessed the link between prior medical conditions and SCZ risk by comparing diagnosis prevalence in cases and controls up to one month before the index date. Table 3 lists twelve conditions with significant differences (FDR < 1%). To address residual confounding and condition overlap, we conducted multivariable regression analysis, incorporating all identified conditions and adjusting for relevant covariates. Results are shown in a forest plot (Fig. 2).

**Figure 2:**
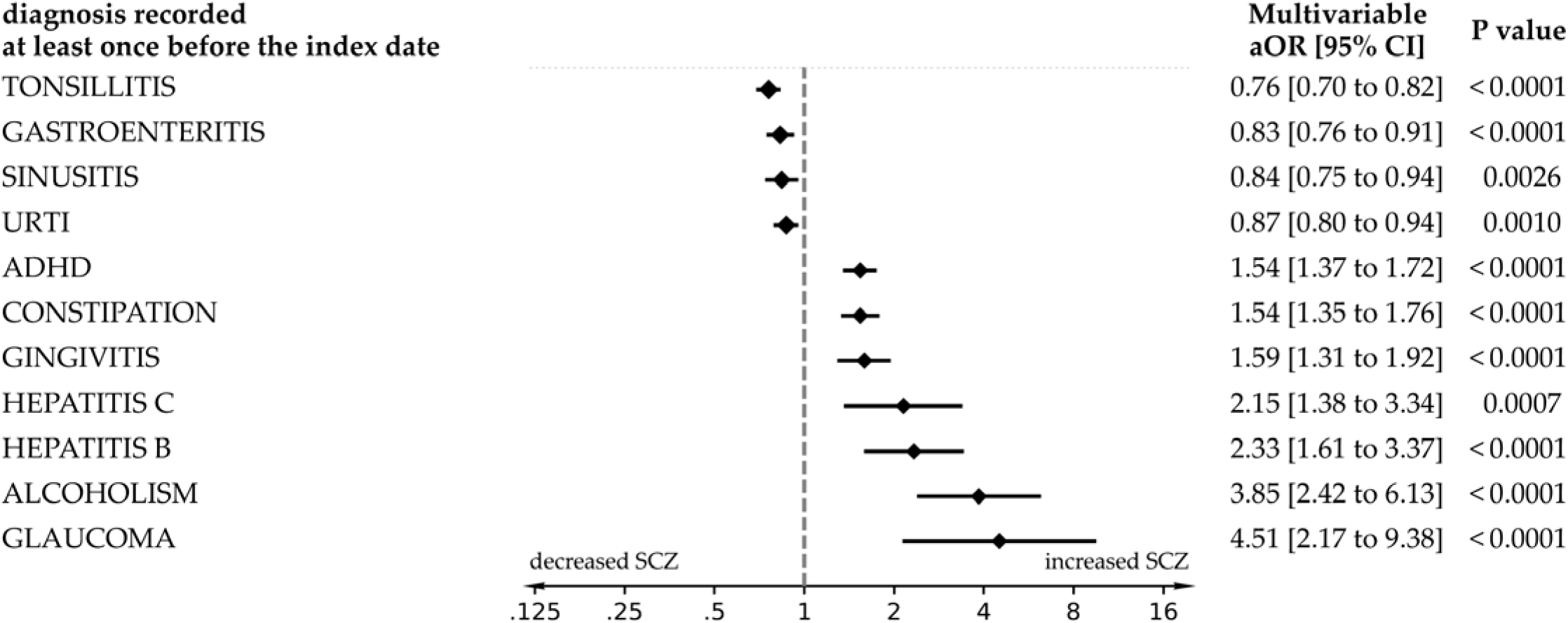
Forest plot of schizophrenia (SCZ) risk according to medical conditions documented in the 20 years preceding the index date. The conditional logistic regression model included all the listed medical conditions and is adjusted for age (years), gender, smoking status, SES category, number of past physician visits, previous pregnancy, and health worker status aOR: Adjusted Odds Ratio; URTI: Upper Respiratory Tract Infection; ADHD: Attention Deficit Hyperactivity Disorder

**Table 3.**
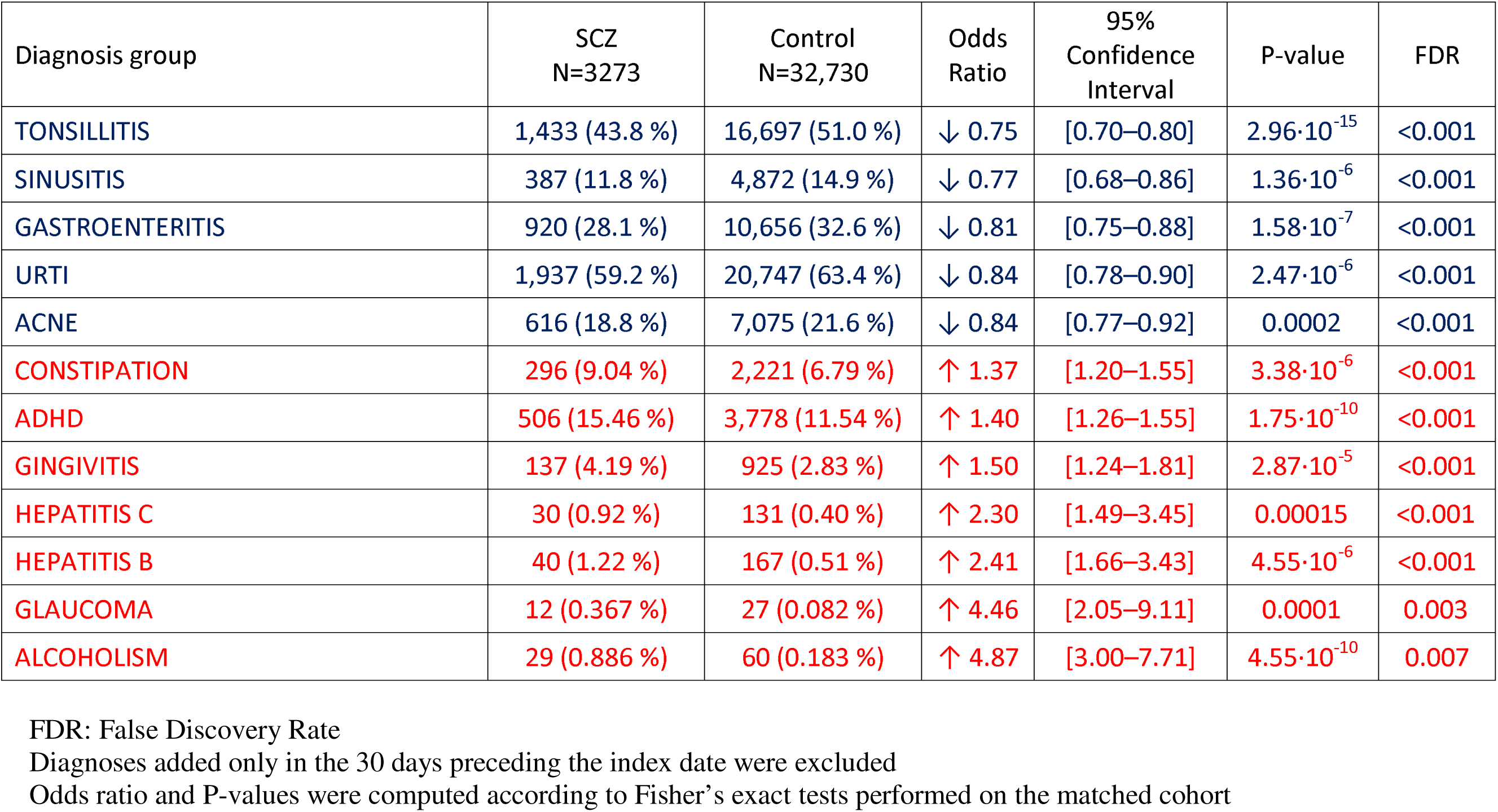
Comparison of diagnoses presence in the 20 years preceding the index date in the schizophrenia (SCZ) and control groups.

| Diagnosis group | SCZ<br>N=3273 | Control<br>N=32,730 | Odds<br>Ratio | 95%<br>Confidence<br>Interval | P-value | FDR |
| --- | --- | --- | --- | --- | --- | --- |
| TONSILLITIS | 1,433 (43.8 %) | 16,697 (51.0 %) | ↓ 0.75 | [0.70–0.80] | $2.96 \cdot 10^{-15}$ | <0.001 |
| SINUSITIS | 387 (11.8 %) | 4,872 (14.9 %) | ↓ 0.77 | [0.68–0.86] | $1.36 \cdot 10^{-6}$ | <0.001 |
| GASTROENTERITIS | 920 (28.1 %) | 10,656 (32.6 %) | ↓ 0.81 | [0.75–0.88] | $1.58 \cdot 10^{-7}$ | <0.001 |
| URTI | 1,937 (59.2 %) | 20,747 (63.4 %) | ↓ 0.84 | [0.78–0.90] | $2.47 \cdot 10^{-6}$ | <0.001 |
| ACNE | 616 (18.8 %) | 7,075 (21.6 %) | ↓ 0.84 | [0.77–0.92] | 0.0002 | <0.001 |
| CONSTIPATION | 296 (9.04 %) | 2,221 (6.79 %) | ↑ 1.37 | [1.20–1.55] | $3.38 \cdot 10^{-6}$ | <0.001 |
| ADHD | 506 (15.46 %) | 3,778 (11.54 %) | ↑ 1.40 | [1.26–1.55] | $1.75 \cdot 10^{-10}$ | <0.001 |
| GINGIVITIS | 137 (4.19 %) | 925 (2.83 %) | ↑ 1.50 | [1.24–1.81] | $2.87 \cdot 10^{-5}$ | <0.001 |
| HEPATITIS C | 30 (0.92 %) | 131 (0.40 %) | ↑ 2.30 | [1.49–3.45] | 0.00015 | <0.001 |
| HEPATITIS B | 40 (1.22 %) | 167 (0.51 %) | ↑ 2.41 | [1.66–3.43] | $4.55 \cdot 10^{-6}$ | <0.001 |
| GLAUCOMA | 12 (0.367 %) | 27 (0.082 %) | ↑ 4.46 | [2.05–9.11] | 0.0001 | 0.003 |
| ALCOHOLISM | 29 (0.886 %) | 60 (0.183 %) | ↑ 4.87 | [3.00–7.71] | $4.55 \cdot 10^{-10}$ | 0.007 |

FDR: False Discovery Rate
Diagnoses added only in the 30 days preceding the index date were excluded
Odds ratio and P-values were computed according to Fisher's exact tests performed on the matched cohort

We observed a significant decrease in SCZ risk among individuals diagnosed with upper respiratory tract infections, including tonsillitis (aOR=0.76 [0.70–0.82]), sinusitis (aOR=0.84 [0.75–0.94]), and other upper respiratory tract infections (URTI) (aOR=0.87 [0.80–0.94]), as well as those with a prior diagnosis of gastroenteritis (aOR=0.83 [0.76–0.91]). Conversely, significant risk increases were noted for constipation (aOR=1.54 [1.35–1.76]), gingivitis (aOR=1.59 [1.31– 1.92]), and attention deficit hyperactivity disorder (ADHD) (aOR=1.54 [1.37–1.72]), aligning with previous studies.^9^ Stronger associations were found with chronic liver conditions, including hepatitis C (aOR=2.15 [1.38–3.34]), hepatitis B (aOR=2.33 [1.61–3.37]), and alcoholism (aOR=3.85 [2.42–6.13]). Glaucoma exhibited the highest risk increase (aOR=4.51 [2.17–9.38]). All associations reflect diagnoses preceding SCZ onset, supporting their temporal precedence.

### Validation in the US based trinetX network

We validated key medication associations from LHS using the U.S. TriNetX network, integrating EHRs from over 69 healthcare organizations and covering >120 million patients. We prospectively assessed SCZ risk linked to specific medications, defining cohorts of individuals initiating treatment between ages 10 and 30, now aged 30–40. These were propensity score-matched to controls from ambulatory medical encounters, excluding prior SCZ diagnoses, with matching for age, gender, ethnicity, BMI, and tobacco use. Results are summarized in Figure 3.

**Figure 3:**
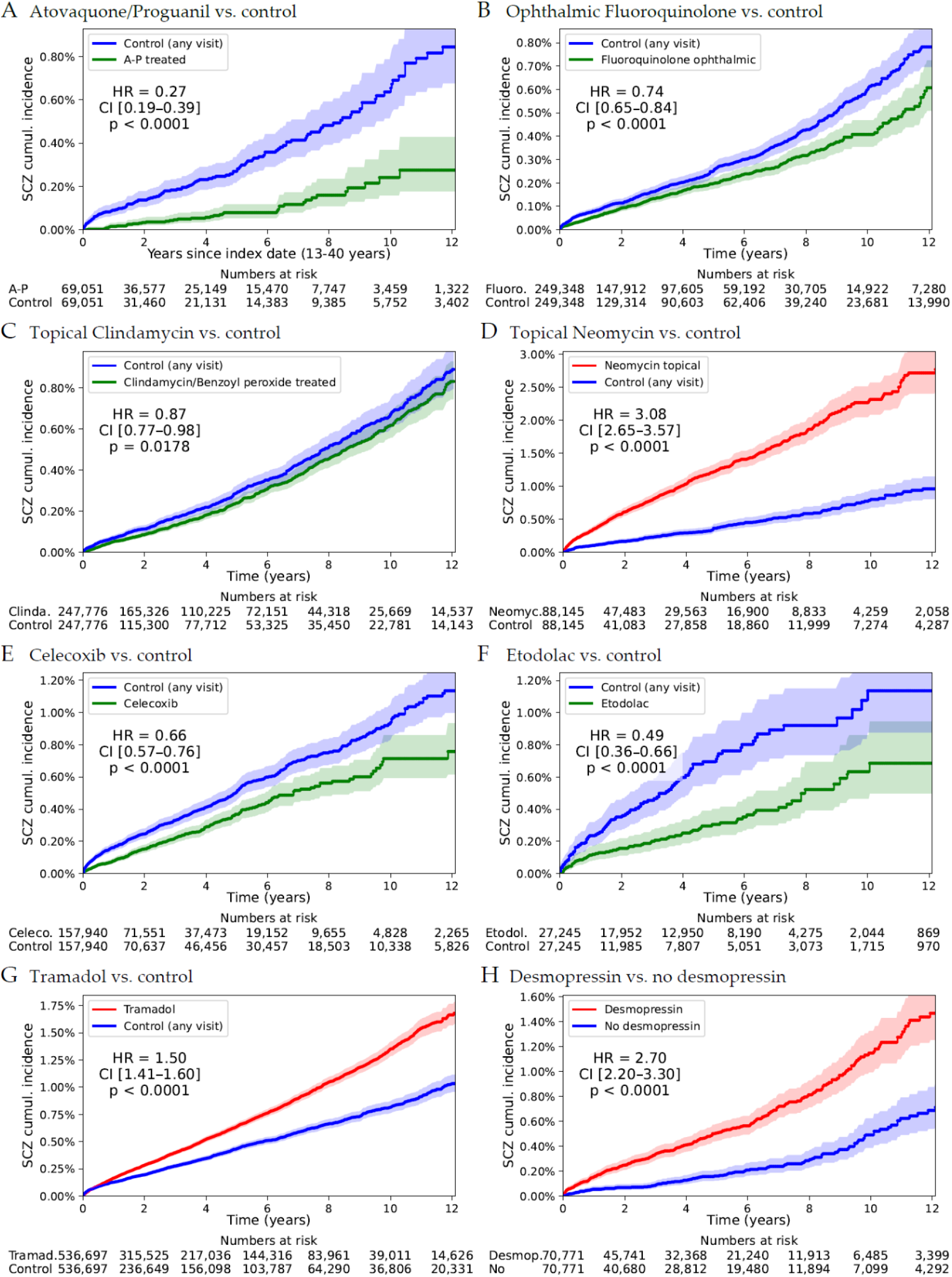
SCZ cumulative incidence in validation cohorts from TriNetX. Medications were tested on a cohort of healthcare organizations (HCOs) within the TriNetX US Collaborative Network, which includes 69 HCOs and a total of 120 million patients. We individuals patients currently aged 13-40 years old and who underwent treatment between the ages of 10 and 30. The index date was defined as the first ambulatory visit occurring at or after medication uptake for the treatment group, or any ambulatory visit for the control group. Patients with a prior diagnosis of schizophrenia (SCZ) before the index date were excluded. Groups were matched using propensity scores, accounting for age, ethnicity, race, smoking status, and, for the desmopressin validation, the presence of urinary incontinence.

For Ato/Pro (Fig. 3A), 69,051 treated patients versus 69,051 controls (from >21 million) showed a Hazard Ratio (HR) of 0.27 [0.19–0.39]. Ophthalmic fluoroquinolones (249,348 patients per group, Fig. 3B) yielded an HR of 0.74 [0.65–0.84], and topical clindamycin (247,776 patients per group, Fig. 3C) an HR of 0.87 [0.77–0.98]. Topical neomycin (88,145 patients per group, Fig. 3D) increased SCZ risk (HR=3.08 [2.65–3.57]). Cox-2 inhibitor celecoxib (157,940 patients per group, Fig. 3E) and partial COX-2 inhibitor etodolac (27,245 patients per group, Fig. 3F) reduced SCZ risk (HR=0.66 [0.57–0.76] and 0.49 [0.36–0.66], respectively). Tramadol (536,697 patients per group, Fig. 3G) increased risk (HR=1.50 [1.41–1.60]). Desmopressin usage (70,771 patients per group, Fig. 3H), matched on urinary incontinence diagnosis, showed a strongest increase (HR=2.70 [2.20–3.30]). All p-values were <0.0001, except clindamycin (p=0.018).

## Discussion

This retrospective population study, performed in a national health provider with over two decades of EHR history, compared 3,273 SCZ patients with 32,730 matched controls. We identified significant associations between past medical treatments, diagnoses, and lab results with SCZ risk, validated in the U.S. TriNetX network (>120 million patients).

Atovaquone/proguanil (Ato/Pro) showed the strongest risk reduction, sustained for over a decade post-treatment. Although only 4 SCZ patients in the LHS cohort had a record of Ato/Pro use, this low number of SCZ cases among treated individuals underscores the strength of the protective association. The adjusted odds ratio remained robust and was further supported by the large-scale TriNetX analysis (n=69,051), addressing potential concerns regarding statistical instability. Protective effects were also found for ophthalmic fluoroquinolones, topical clindamycin, and NSAIDs (notably COX-2 inhibitors). Conversely, neomycin, tramadol, and desmopressin increased SCZ risk. Conditions like chronic liver disease, constipation, and glaucoma were risk factors, while gastroenteritis and upper respiratory infections were protective.

### Atovaquone/Proguanil and clindamycin

Atovaquone/proguanil (Ato/Pro) was linked to the strongest reduction in SCZ risk (aOR=0.26, LHS; HR=0.27, P<10 ²², TriNetX). Primarily prescribed for malaria prophylaxis in travelers to endemic regions, its effect on SCZ risk is unlikely to relate to malaria, as the disease is non-endemic in Israel and the U.S. Ato/Pro’s components target protozoan-specific pathways absent in human cells: atovaquone inhibits the mitochondrial electron transport chain, and proguanil, via its metabolite cycloguanil, blocks dihydrofolate reductase (DHFR), disrupting DNA synthesis. The sustained SCZ risk reduction - persisting over a decade post-treatment - suggests elimination of a pathogen involved in SCZ pathogenesis rather than a direct neurophysiological effect.

The most plausible explanation is the clearance of *T. gondii*, a parasite consistently linked to SCZ.^10,11^ A meta-analysis of 38 studies across 14 countries found a strong association between *T. gondii* seropositivity and SCZ.^3^ Atovaquone, effective against *T. gondii*, is often used in toxoplasmosis treatment, albeit at higher doses than for malaria prophylaxis.

*T. gondii* encysts in dopamine-rich brain regions, producing a tyrosine hydroxylase-like enzyme that boosts dopamine synthesis. Some antipsychotics possess anti-*T. gondii* properties, which may explain their efficacy in infected patients.^12^

The abrupt elimination of *T. gondii* by Ato/Pro may disrupt dopamine regulation. This, combined with the immune response triggered by parasite clearance, could induce neuroinflammation, further affecting dopamine pathways, potentially explaining rare reports of psychotic symptoms following treatment onset. ^13^

Our analysis also identified topical clindamycin - an effective anti-*T. gondii* agent*^14,15^* - was associated with a significantly reduced risk of SCZ, both when used alone and in combination with benzoyl peroxide. We propose that its prolonged use in acne treatment may deplete *T. gondii* populations on the skin. Additionally, repeated hand contact during application could interrupt parasite transmission through self-inoculation, thereby reducing T. gondii’s ability to spread between sites.

In contrast, our analysis identified an increased risk of SCZ associated with several topical antibiotics, including fusidic acid, chloramphenicol, gentamicin, and mupirocin. Notably, two neomycin-based compounds exhibited particularly strong associations with elevated SCZ risk, with aOR of 1.17 and 1.55, respectively. This increased risk was further confirmed in the TriNetX cohort, where 87,207 individuals who received topical neomycin had a 3-fold higher risk of SCZ compared with matched controls.

These antibiotics are known to disrupt the commensal skin microbiota, which normally inhibits pathogen colonization, potentially creating an environment conducive to opportunistic pathogens such as *T. gondii*.^16^ Remarkably, a recent study reported a persistent reduction in commensal *Staphylococcus* species - important for protecting the skin against invasive pathogens - following treatment with a neomycin-containing ointment.^17^ Consistent with this, in our analysis, the highest observed increase in SCZ risk was linked to an ointment-based neomycin treatment.

### Ophthalmic antimicrobial medications and SCZ risk

Our study identified a reduced risk of SCZ for two ophthalmic antibiotics. Use of the fluoroquinolone moxifloxacin was associated with a 47% reduction in SCZ risk (aOR=0.53), while an oxytetracycline-based compound had a milder protective effect (aOR=0.61). We validated this finding in the TriNetX cohort, where treatment with ophthalmic fluoroquinolones across 248,881 patients also showed SCZ risk reduction compared to their matched controls (HR=0.74).

Fluoroquinolones exert their antimicrobial effects by inhibiting DNA gyrase and topoisomerase IV, enzymes essential for bacterial DNA replication. Interestingly, studies show they also interfere with *T. gondii* replication by either targeting the apicoplast, a plastid crucial for parasite survival, or disrupting mitochondrial function, thereby impairing parasite proliferation.^18,19^ A similar inhibitory effect of *T. gondii* was also observed for oxytetracycline,^20^ which inhibits the apicoplast 30S ribosomal unit, blocking translation.

Given the high prevalence of *T. gondii* in individuals with SCZ, the observation that five compounds with known anti-*T. gondii* activity - notably Ato/Pro, two clindamycin formulations, moxifloxacin, and oxytetracycline - are associated with the highest reductions in SCZ risk in two distinct cohorts, strongly supports a causal role for the parasite in SCZ pathology.

The susceptibility of the SCZ-associated pathogen to ophthalmic antibiotics suggests that the eye may act as an entry point to the brain. *T. gondii* is known to infect the eye, causing ocular toxoplasmosis^21^, which commonly presents as retinochoroiditis, the leading cause of posterior uveitis worldwide. Alternatively, sterilization of the oronasal cavity or paranasal sinuses - where ophthalmic antibiotics may accumulate via lacrimal duct drainage - could eliminate pathogens at these sites, thereby hindering their spread to the brain.

Further supporting this hypothesis is our finding that glaucoma was the most significant medical condition associated with SCZ occurrence (aOR=4.51), a finding consistent with previous studies.^22^ Here, this association was evident even before SCZ diagnosis, suggesting that glaucoma and SCZ may share a common risk factor. Although excessive ocular pressure is the primary mechanism underlying optic nerve degeneration in glaucoma, its early onset before SCZ suggests the involvement of an additional factor, possibly a direct *T. gondii*-mediated insult to optic nerve cells.

### Upper respiratory infections and SCZ risk

Our analysis shows that medical conditions that trigger an increased immune response in the upper respiratory system are associated with a reduced risk of SCZ, particularly sinusitis, tonsillitis, and URTIs.

A common trait of these infections is that they induce a strong localized immune response, including increased secretion of secretory IgA, activation of macrophages and neutrophils, and upregulation of pro-inflammatory cytokines, all of which may help eliminate *T. gondii* from the oronasal mucosa. Additionally, pathogen competition and microbiome alterations caused by bacterial and viral infections could create an unfavorable environment for *T. gondii* adherence and invasion. Frequent epithelial turnover due to inflammatory damage may further prevent parasite persistence, while increased mucus secretion and ciliary clearance could physically remove *T. gondii* from the mucosal surface.

Conversely, we identified that gingivitis was significantly associated with increased SCZ risk (aOR=1.59). Gingivitis is a condition characterized by inflammation of the gingiva caused by pathogens residing in dental plaque. Gingivitis could both facilitate *T. gondii* infection by weakening the oral mucosal barrier and serve as an indicator of underlying oral toxoplasmosis, especially in individuals with recurrent or severe periodontal disease.

Moreover, periodontal inflammation may also promote the recruitment of monocytes and macrophages, which *T. gondii* can use as Trojan horses for dissemination to other tissues, including the brain.^23^

Interestingly, our study identified a moderate protective effect for three antibiotics effective against oral bacteria involved in gingivitis and periodontal infections: phenoxymethylpenicillin (aOR=0.72), amoxicillin/clavulanate (aOR=0.86), and azithromycin (aOR=0.91). Depletion of pathogenic bacteria from oral and upper respiratory sites may explain how they might contribute to reduced risk of SCZ.

### The gastrointestinal tract and SCZ risk

Our analysis identified two gastrointestinal transit disturbances as significantly affecting SCZ risk, in opposite directions, strongly suggesting a role for the gut in SCZ pathogenesis: constipation was associated with a notable increase in SCZ risk (aOR=1.54), whereas gastroenteritis was linked to a significant risk reduction (aOR=0.83).

The gut may provide a more favorable environment for *T. gondii* replication than the CNS or skin. In the CNS, neurons – being terminally differentiated cells with low turnover rates – create a stable environment that favors the parasite’s transition into the bradyzoite latent state within tissue cysts, minimizing immune detection and limiting active replication. In the skin, *T. gondii* encounters physical and immune barriers, as well as low metabolic activity and limited cell turnover, further restricting its invasion and proliferation potential.

In contrast, the intestinal epithelium undergoes continuous and rapid renewal, with enterocytes being replaced approximately every 4 to 5 days. This high turnover rate provides *T. gondii* with ample opportunities to invade new cells, sustaining its replication cycles. In primary infections, *T. gondii* usually invades enterocytes in the small intestine, where the parasite replicates, as tachyzoites, before disseminating to other tissues. Furthermore, the gut-associated lymphoid tissue (GALT) in the colon is designed to tolerate a wide range of antigens, including commensal microbiota, while maintaining immune homeostasis. This tolerogenic environment may allow *T. gondii* to evade immediate immune detection, facilitating its persistence and replication within intestinal cells. Additionally, the presence of immune cells in the lamina propria can aid in the parasite’s systemic dissemination once it breaches the epithelial barrier.

Altered intestinal transit may further influence *T. gondii* replication and persistence. Reduced intestinal motility, such as in constipation, may provide an even more favorable environment by prolonging parasite contact with the intestinal epithelium, enhancing invasion and replication within enterocytes. Slow transit may also promote microbial fermentation and gut microbiome alterations that reduce immune surveillance, supporting local parasite persistence, explaining the increased risk observed in constipation.

In contrast, gastroenteritis is associated with increased intestinal transit, limiting the time available for *T. gondii* tachyzoites or bradyzoites to attach to and invade host cells. Moreover, increased peristalsis and rapid fluid loss may further facilitate the physical expulsion of parasites from the gut, potentially explaining how gastroenteritis reduces SCZ risk.

### Impaired liver function and SCZ risk

Four of the medical conditions associated with strong increases in SCZ risk in our analysis were linked to liver dysfunction: alcoholism (aOR=3.85), hepatitis C (aOR=2.15), and hepatitis B (aOR=2.33), consistent with previous findings^24,25^ but here shown to precede SCZ onset. Conversely, the hepatitis A vaccine was associated with a strong decrease in SCZ risk (aOR=0.31). Introduced into Israel’s routine immunization program in 1999,^26^ this vaccine prevents hepatitis A, a frequent cause of transient liver dysfunction, often asymptomatic, in children and adults.

Further evidence of hepatic involvement in SCZ is reflected in disproportionate reductions in total T3 relative to free T3 in SCZ patients (SMD=-0.409 vs. -0.049, respectively), indicating reduced thyroid-binding globulin (TBG) synthesis, a marker of impaired hepatic protein production. The liver plays a central role in immune surveillance and pathogen clearance, including defense against *T. gondii*. It filters pathogens from the bloodstream, activates immune responses via Kupffer and NK cells, and produces complement proteins crucial for parasite recognition and clearance. Liver dysfunction compromises these defenses, potentially weakening *T. gondii* containment and facilitating its systemic spread.

Thus, liver impairment - whether due to viral hepatitis or alcoholism - may increase susceptibility to *T. gondii* infection, contributing to neuroinflammation and SCZ risk. These findings highlight the importance of liver function in immune regulation and suggest a potential role in SCZ pathogenesis.

### NSAIDs reduce SCZ risk

In our study, treatment with several nonsteroidal anti-inflammatory drugs (NSAIDs), including etoricoxib (aOR=0.73), etodolac (aOR=0.73), and ibuprofen (aOR=0.77), was significantly associated with reduced SCZ risk. This finding was further supported by an analysis in the TriNetX database, where celecoxib use was linked to a strong reduction in SCZ risk, reinforcing the potential protective role of COX-2 inhibitors.

Neuroinflammation is implicated in SCZ pathogenesis,^27^ with elevated pro-inflammatory cytokines^28^ and microglial activation^29^ contributing to synaptic pruning and neuronal damage.^29^ COX-2 inhibitors may mitigate these effects,^30^ with adjunctive NSAID treatment previously shown to improve psychotic symptoms. Moreover, neuroinflammation may compromise the blood-brain barrier (BBB)^31^, facilitating *T. gondii* invasion via infected immune cells. NSAIDs, by stabilizing the BBB^32^ and suppressing microglial activation, could help prevent parasite entry and reduce neuronal damage, further supporting their potential role in SCZ prevention.

### Tramadol increases SCZ risk

In contrast to NSAIDs, tramadol use was significantly associated with increased SCZ risk (aOR=1.50 for combined forms with paracetamol, 1.62 for tramadol alone). This association was confirmed in the TriNetX cohort, where tramadol recipients had a higher SCZ risk (HR=1.50), consistent with prior links between opioid use disorder and SCZ progression.

Tramadol’s inhibitory effect on intestinal peristalsis may promote *T. gondii* replication by prolonging parasite contact with the intestinal epithelium. Additionally, opioids suppress immune function,^33^ potentially allowing greater *T. gondii* CNS infiltration, and increase BBB permeability by disrupting tight junctions, facilitating parasite entry. Opioids also exacerbate neuroinflammation and glial dysregulation,^34^ which may contribute to SCZ pathogenesis.

These findings highlight the need for cautious opioid prescribing and close monitoring in individuals at risk for SCZ.

### Desmopressin increases SCZ risk

Desmopressin, a vasopressin analog prescribed for nocturnal enuresis, was the medication associated with the strongest increase in SCZ risk (aOR=2.04). A validation study in TriNetX (n=70,775 per group) confirmed this finding, with desmopressin use remaining significantly associated with SCZ risk (HR=2.70, *P* < 0.0001) even after propensity matching for urinary incontinence.

Vasopressin-related genes have been implicated in SCZ,^35^ with upregulation of *AVP* and *AVPR1a* in SCZ patients.^36^ AVP also modulates blood-brain barrier (BBB) permeability and immune cell trafficking,^37^ potentially facilitating *T. gondii* neuroinvasion. Moreover, AVP overstimulation triggers microglial activation, contributing to neuroinflammation and neurotransmitter dysregulation in SCZ.

These findings highlight the need for caution when prescribing vasopressin agonists to individuals at risk for SCZ.

### Laboratory tests and SCZ risk

At disease onset (Table 1), SCZ patients had significantly lower levels of T3, vitamin D, and folic acid compared to controls. T3 deficiency (free T3 <3.5 pmol/L, OR=1.8) and elevated TSH (>4.5 mIU/L, OR=2.0) suggest increased rates of subclinical or clinical hypothyroidism in SCZ individuals. Thyroid hormones regulate microglial activity and blood-brain barrier (BBB) integrity, and lower T3 levels may impair pathogen clearance and increase BBB permeability, facilitating *T. gondii* neuroinvasion.

Vitamin D insufficiency (<30 ng/mL, OR=1.38) was also more prevalent in SCZ patients^5^, consistent with its role in antimicrobial defense.^38–40^ Vitamin D stimulates macrophages and dendritic cells to produce antiparasitic peptides and enhances IFN-γ production, crucial for *T. gondii* control. Deficiency may weaken immune defenses, promoting chronic infection.^41,42^

Folic acid deficiency (OR=1.41) was similarly associated with SCZ. Folate supports T-cell–mediated immunity and microglial function, both essential for *T. gondii* elimination. Low levels may impair immune responses, increasing susceptibility to persistent infection.

These findings highlight potential immune-mediated mechanisms linking micronutrient deficiencies to SCZ risk.

## Conclusion

This large-scale, retrospective study, spanning 20 years within a national health provider and validated in the TriNetX U.S. network, identified significant associations between past medical treatments and schizophrenia (SCZ) risk, revealing both novel protective and risk factors. Treatment with atovaquone/proguanil (Ato/Pro), topical clindamycin, and ophthalmic fluoroquinolones - agents known to be effective against *Toxoplasma gondii* - was strongly associated with a sustained reduction in SCZ risk, persisting for over a decade after exposure. This pattern, together with the consistently elevated *T. gondii* seroprevalence observed in SCZ patients as established by multiple studies, supports a potential etiological role for this pathogen. Use of NSAIDs, particularly COX-2 inhibitors, was also linked to reduced SCZ risk, reinforcing the contribution of neuroinflammation to disease pathogenesis and pointing to potential therapeutic pathways.

While these findings are compelling, they should be interpreted with caution. Despite matching on key demographic, socioeconomic, and healthcare utilization variables, residual confounding may still influence associations, as is inherent to all observational studies. Importantly, we did not directly measure *T. gondii* infection status in our cohorts. Our mechanistic interpretations are grounded in the well-established serological association between *T. gondii* and SCZ shown in multiple studies and the observation that medications targeting this parasite, notably Ato/Pro, clindamycin, and fluoroquinolones, were consistently linked to sustained reductions in SCZ risk in the two cohorts studied.

Taken together, our results support a unifying hypothesis in which chronic *T. gondii* infection-enabled by immune dysfunction, impaired liver function, microbiota disruption, and neuroinflammation - may contribute to SCZ development. Future studies should directly investigate *T. gondii* presence in biological samples from SCZ patients. Targeted elimination strategies - combining antiparasitic agents, COX-2 inhibitors, and dopamine-stabilizing therapies - warrant investigation in prospective trials to assess their potential in SCZ prevention and treatment.

## Data Availability

Access to patients data is limited to researchers approved by the Institutional Review Board.

## Author contributions

Conceptualization: AI, EMe, Ema

Methodology: AI, Eme

Investigation: AI, SI, JS, Eme

Writing – original draft: AI

Writing – review & editing: AI, AW, SI, JS, SA, SV, EMa, EMe

## Materials & Correspondence

Correspondence and requests for materials should be addressed to AI

## Abbreviations

ADHD: Attention deficit hyperactivity disorder
AVP: Vasopressin
AVPR1a: Vasopressin receptor 1A
aOR: adjusted Odds Ratio
BMI: Body Mass Index
BP: Blood pressure
CNS: Central nervous system
COX-2: Cyclooxygenase-2
EHR: Electronic health records
FDR: False Discovery Rate
HCO: Healthcare organizations
HR: Hazard Ratio
ICD-9: International Classification of Diseases, Ninth Revision
IM: Intramuscular
LHS: Leumit Health Services
OR: Odds Ratio
PO: Per os
PSM: Propensity score matching
SCZ: Schizophrenia
SES: Socioeconomic status
SMD: Standardized Mean Difference
T3: Triiodothyronine
TBG: Thyroid-binding globulin
TSH: Thyroid-stimulating hormone
URTI: Upper Respiratory Tract Infection

